# High SARS-CoV-2 Seroprevalence in Children and Adults in the Austrian Ski Resort Ischgl

**DOI:** 10.1101/2020.08.20.20178533

**Authors:** Ludwig Knabl, Tanmay Mitra, Janine Kimpel, Annika Rössler, André Volland, Andreas Walser, Hanno Ulmer, Lisa Pipperger, Sebastian C. Binder, Lydia Riepler, Katie Bates, Arnab Bandyopadhyay, Marta Schips, Mrinalini Ranjan, Barbara Falkensammer, Wegene Borena, Michael Meyer-Hermann, Dorothee von Laer

**Author notes:** Corresponding authors: Michael Meyer-Hermann, PhD, Department of Systems Immunology and Braunschweig Integrated Centre of Systems Biology, Helmholtz Centre for Infection Research, Braunschweig, Germany E-Mail Dorothee von Laer, MD, Institute of Virology, Medical University of Innsbruck, Schöpfstraße 41, AT-6020 Innsbruck, Austria. These authors contributed equally to this work (Ludwig Knabl, Tanmay Mitra, Janine Kimpel, Wegene Borena, Michael Meyer-Hermann, Dorothee von Laer).

## Abstract

**Background:** Early March 2020, a SARS-CoV-2 outbreak in the ski resort Ischgl in Austria initiated the spread of SARS-CoV-2 throughout Austria and Northern Europe.

**Methods:** Between April 21 and 27, a cross-sectional epidemiologic study targeting the full population of Ischgl (n = 1867), of which 79 % could be included (n = 1473, incl. 214 children), was performed. For each individual, the study involved a SARS-CoV-2 PCR, antibody testing and structured questionnaires. A mathematical model was used to help understand the influence of the determined seroprevalence on virus transmission.

**Findings:** The seroprevalence was 42.4% (95% CI 39.8–44.7). Individuals under 18 showed a significantly lower seroprevalence of 27.1% (95% CI 21.3–33.6) than adults (45%; 95% CI 42.2–47.7; OR of 0.455, 95% CI 0.356–0.682, p< 0.001). Of the seropositive individuals, 83.7% had not been diagnosed to have had SARS-CoV-2 infection previously. The clinical course was generally mild. Over the previous two months, two COVID-19-related deaths had been recorded, corresponding to an infection fatality rate (IFR) of 0.25% (95% CI 0.03–0.91). Only 8 (0.5 %) individuals were newly diagnosed to be infected with SARS-CoV-2 during this study.

**Interpretation:** Ischgl was hit early and hard by SARS-CoV-2 leading to a high local seroprevalence of 42.4%, which was lower in individuals below the age of 18 than in adults. Mathematical modeling suggests that a drastic decline of newly infected individuals in Ischgl by the end of April occured due to the dual impact from the non-pharmacological interventions (NPIs) and a significant immunization of the Ischgl population.

**Funding:** Helmholtz Association, European Union’s Horizon 2020 research and innovation program, German Research Foundation (DFG), state Tyrol.

## Introduction

Since December, the severe acute respiratory syndrome coronavirus 2 (SARS-CoV-2) has spread from Wuhan, China, to all continents causing a severe pandemic.^1–3^ Globally, as of end of July, 2020, there were over 17 million confirmed cases of COVID-19, including over 600,000 deaths, reported to WHO.^4^ In Austria, the first cases were detected on February 25^th^, 2020 and until early March, only few limited outbreaks were observed. Then a super-spreading event occurred in an après-ski bar in the ski resort Ischgl leading to an explosive local outbreak associated with the spread of the virus throughout Austria and to many other European countries and worldwide.^5,6^ Since then, epidemiological cluster analyses performed by the Austrian Agency for Health and Food Safety (AGES) found that by April 20^th^ up to 40% of SARS-CoV-2 infections in Austria could be traced back to the epicenter Ischgl.^7^

The bar was closed on March 8 and all après-ski bars in Ischgl on the 10^th^. After the tourists and most guest workers had left, the valley, in which Ischgl lies, was quarantined on March 13 and remained completely isolated for 6 weeks before our study started. In this isolated population, we determined sero- and viruspositivity, modeled the effect of the high seropositivity on virus transmission and analyzed the clinical course, hospitalization and fatality rate, and percentage of non-diagnosed cases.

## Methods

### Study population, study design and recruitment

The ethical committee of the Medical University of Innsbruck approved the study (EC numbers: 1100/2020 and 1111/2020), which took place between April 21 and 27, 2020. This cross-sectional epidemiological survey targeted all residents of Ischgl/Tyrol irrespective of age and gender. At the time of investigation, Ischgl had a population size of 1,867 individuals - 1,617 with their main residence in Ischgl and 250 seasonal immigrant workers - living in 582 different households.^8^ The number of seasonal workers is a value estimated by the local authorities. Most of the over 2000 workers present in Ischgl early March left before or directly after the quarantine was initiated without signing out. All households in Ischgl were contacted by a postal invitation to attend the study center at a specific time for each household, whereby 1,534 individuals from 478 households participated, corresponding to a participation rate of 82.7% and 63.5% of the adult and pediatric population, respectively (Figure S1). As the study was anonymous and some individuals did not visit the study center together with the rest of the household members, 106 individuals could not be assigned to a household. After exclusion of 61 study participants (due to missing of one of the biosamples, i.e. blood sampling was not possible in 2 children and 59 adults refused swab sampling), 1,473 (79% of the Ischgl population) were included into statistical analysis. Structured questionnaires were used to collect data on age, household affiliation, occurrence of physical symptoms after February 1, 2020, and previous SARS-CoV-2 PCR results. The basic characteristics of the study participants are given in Table 1. The participants were screened for anti-SARS-CoV-2 antibodies as well as for SARS-CoV-2 virus in nasopharyngeal swabs by 3 immunoassays for binding antibodies, a neutralization antibody assay (nAb-assay) and an RT-PCR, respectively.

**Table 1.** Baseline characteristics of study participants.

| Variables | Value |
| --- | --- |
| **Number of participants, n (%)** | 1473 (100) |
| Male | 712 (48.3) |
| Female | 759 (51.5) |
| Diverse | 2 (0.1) |
| **Age (years), mean (SD), median** |  |
| Male | 40.3 (19.5) 40 |
| Female | 40.9 (20.1) 40 |
| Children (<18) | 10.2 (4.9) 10.5 |
| Adults (≥ 18) | 45.8 (16.5) 45 |
| **Age (years) distribution, n (%)** |  |
| <6 | 52 (3.5) |
| 6-9 | 49 (3.3) |
| 10-13 | 57 (3.9) |
| 14-17 | 56 (3.8) |
| 18-24 | 124 (8.4) |
| 25-34 | 256 (17.4) |
| 35-44 | 239 (16.2) |
| 45-54 | 238 (16.2) |
| 55-64 | 225 (15.3) |
| 65-74 | 117 (7.9) |
| ≥ 75 | 60 (4.1) |

**B. Clinical and laboratory data**
| Variable | Adults | Children |
| --- | --- | --- |
| **Reported hospitalization due to SARS-CoV-2, n (%)** |  |  |
| Male | 7 (1.2) | 0 |
| Female | 2 (0.3) | 0 |
| Diverse | 0 | n.a. |
| **PCR self-reported positive, n (%)** |  |  |
| Male | 45 (7.4) | 2 (1.9) |
| Female | 54 (8.3) | 4 (3.6) |
| Diverse | 0 | n.a. |
| **PCR tested ad study site positive\*, n (%)** |  |  |
| Male | 4 (0.7) | 2 (1.9) |
| Female | 2 (0.3) | 1 (0.9) |
| Diverse | 0 | n.a. |
| **Seroprevalence (2 of 4 antibody tests positive), n (%)** |  |  |
| Male | 284 (46.7) | 26 (25.0) |
| Female | 281 (43.3) | 32 (29.1) |
| Diverse | 1 (50) | n.a. |
| *\* one of the nine study-site-PCR-positive participants was tested SARS-CoV-2-PCR positive before (on 23.03.2020)* |  |  |

### SARS-CoV-2-PCR, anti-SARS-CoV-2-antibody tests

RT-PCR for the detection of SARS-CoV-2 was performed using the RealStar^®^ SARS-CoV-2 RT-PCR kit 1.0 (Altona Diagnostics GmbH, Hamburg, Germany). Ct values below 40 were rated as positive and confirmed by a second PCR. The district administration provided results of SARS-CoV-2 PCR tests in Ischgl, which had been performed between March 1 and April 20, 2020.

Participants’ sera were screened for anti-SARS-CoV-2-S1-protein IgA and IgG positivity by a commercially available anti-SARS-CoV-2-IgA and -IgG ELISA (Euroimmun, Lübeck, Germany), respectively, and for anti-SARS-CoV-2-N-protein IgG (anti-N IgG) with the Abbott SARS-CoV-2 IgG immunoassay (Abbott, Illinois, USA). Borderline values in the Euroimmun IgG ELISA were rated positive.

### Neutralizing antibody-assay

Neutralizing antibody (nAb)-assays using highly SARS-CoV-2 susceptible cells^9^ were performed as described in detail in the supplement. We tested all 50 anti-S IgG+/anti-N IgG+ positive children, 148 randomly selected currently PCR-negative adults that were positive for anti-S IgG as well as anti-N-IgG, all PCR-negative subjects with discrepant antibody results (n = 38 for anti-S IgG+/anti-N IgG- and n = 36 for anti-S IgG-/anti-N IgG+) or with solely anti-S IgA positive ELISA results (n = 26). Additionally, all 9 currently PCR positive patients were analyzed (Table 2, Figure S4). Plasma samples from 10 blood donors from early 2019 served as negative controls.

**Table 2.** Determining true antibody positivity

|  | Anti-S IgG +<br>Anti-N IgG+ | Anti-S IgG +<br>Anti-N IgG- | Anti-S IgG -<br>Anti-N IgG+ | Anti-S<br>IgA only* | 3 ELISAs<br>negative<br>and PCR<br>positive** | 3 ELISAs<br>negative<br>and PCR<br>negative** |
| --- | --- | --- | --- | --- | --- | --- |
| **Total**<br>**N=1473 (%)** | 544<br>(36.9) | 38<br>(2.6) | 36<br>(2.4) | 26<br>(1.7) | 6<br>(0.4) | 823<br>(55.9) |
| **Neutralisation**<br>**Assay >1:4** | 201/201<sup>&</sup><br>(100) | 32/37<sup>#</sup><br>(88.5) | 36/36<br>(100) | 10/26<br>(38.5) | 1/3<br>(33.3) | n.a. |
| **Rate as positive**<br>**n=624 (%)** | 544 (87.2) | 34 (5.4) | 36 (5.8) | 10 (1.6) | 0 (0.0) | 0 (0.0) |
| **Reported PCR**<br>**positive**<br>**n= 105 (%)** | 95(90.5) | 3(2.9) | 4 (3.8) | 0 (0.0) | 3 (2.9) | 0(0.0) |
| **New PCR-**<br>**confirmed cases**<br>**n= 8<sup>†</sup> (%)** | 2 (25) | 1 (12.5) | 0 (0.0) | 2 (25) | 3 (37.5) | 0 (0.0) |

<sup>&</sup>Not all Anti-S IgG + Anti-N IgG+ positive samples were tested in the neutralization assay,
<sup>#</sup> No sample left for neutralizing antibody assay for 1 patient
\*Anti-S IgA available for n=975 participants, <sup>†</sup>One person excluded as PCR positivity already known (included in the reported PCR positive group)
S= spike protein. N= nucleocapsid protein, n.a. = not applicable

As the immunoassays can produce false positive results, they are generally confirmed by a neutralizing antibody assay. However, out of 400 Sera from 2019, no serum was found to be positive in both immunoassays used here (manuscript in preparation). Thus, the combination of both antibody assays, requiring both to be positive for a final positive result, had a specificity of 100%. Therefore, all sera were tested in two different antibody assays and samples that were positive in both anti-S IgG and anti-N-IgG antibody assays were not all retested by the neutralizing antibody assay. To maximize sensitivity, all antibody discrepant sera were tested in the neutralizing antibody assay. Thereby a sensitivity of nearly 100% for the presence of antibodies to SARS-CoV-2 is expected to be achieved. Reported performances of the antibody assays used have been described in detail previously^10^.

For the maximally four different methods for anti-SARS-CoV-2 antibody detection, the following finding constellations were classified as seropositive:

I. Positive result in anti-S1-protein IgG ELISA AND anti-N-protein IgG immunoassay.
II. Positive result in anti-S1-protein IgG ELISA OR anti-N-protein IgG immunoassay and a positive result in the nAb-assay.
III. Positive result in anti-S1-protein IgA ELISA AND positive result in nAb-Assay.

### Statistical and mathematical analysis

Demographic characteristics were tabulated using descriptive statistics including the calculation of means ± standard deviations for continuous measures and numbers (%) for categorical measures. 95% confidence intervals for crude prevalence estimates (self-reported PCR testing, seroprevalence, hospitalization and infection fatality rate), overall and stratified by sex and adults/children, were computed by the method of Clopper and Pearson^11^. General estimating equation (GEE) models according to Zeger and Liang were employed to model associations of self-reported symptoms with seropositivity outcomes, taking into account the possible interdependencies of study participants living in the same household^12^. The age- and sex adjusted GEE models were specified with a logistic link function, an exchangeable correlation structure and robust standard errors to estimate odds ratios (OR) and 95% confidence intervals (CI). To prevent confounding by flu symptoms, self-reported symptoms occurring during peak influenca season (before February 23) were excluded from the GEE analysis.

A household analysis of all households was performed as described in detail in the supplement.

The SARS-CoV-2 outbreak in Ischgl was modeled using a compartmental model of infection epidemics based on ordinary differential equations (ODEs), specifically adapted to the biological features and clinical course associated with SARS-CoV-2 infection (see Supplementary Information).

## Results

### Seroprevalence

From a total of approximately 1867 people living in Ischgl, 1473 individuals living in 478 households including all age groups were tested for antibodies to SARS-CoV-2 and for SARS-CoV-2 virus RNA in nasopharyngeal swabs (Figure S1). This included 101 children under the age of 10 and 113 participants between 10 and 17 (Table 1). A total of 624 study participants were seropositive in at least 2 of maximally 4 antibody tests performed (3 binding antibody assays + neutralizing antibody test) corresponding to a seroprevalence of 42.4 % (95% CI 39.8–44.7, Table 2). The IgG values for the anti-SARS-CoV-2 S protein antibodies (Euroimmun ELISA) correlated well with the anti-N IgG levels (Abbott immunoassay) (Figure S2, R = 0.8). All sera that were positive in only one of the two assays were tested for neutralizing antibodies (Table 2, Figure S3). The values in the anti-S protein IgG ELISA correlated well with the neutralizing antibody titers, R = 0.6, p< 0.001 (Figure S4). The seroprevalence was significantly lower in children and adolescents under 18 than in adults, with 27.1% (95% CI (21.0–33.6)) and 45% (95% CI (42.2–47.7)), respectively (OR of 0.455, 95% CI 0.356–0.682, p< 0.001; Figure 1A). The seroprevalence in women 41.2% (37.5–44.9) did not differ significantly from men 43.5% (39.5–47.3) (OR 0.91 (0.74–1.12), p = 0.37; Figure 1A).

**Figure 1.**
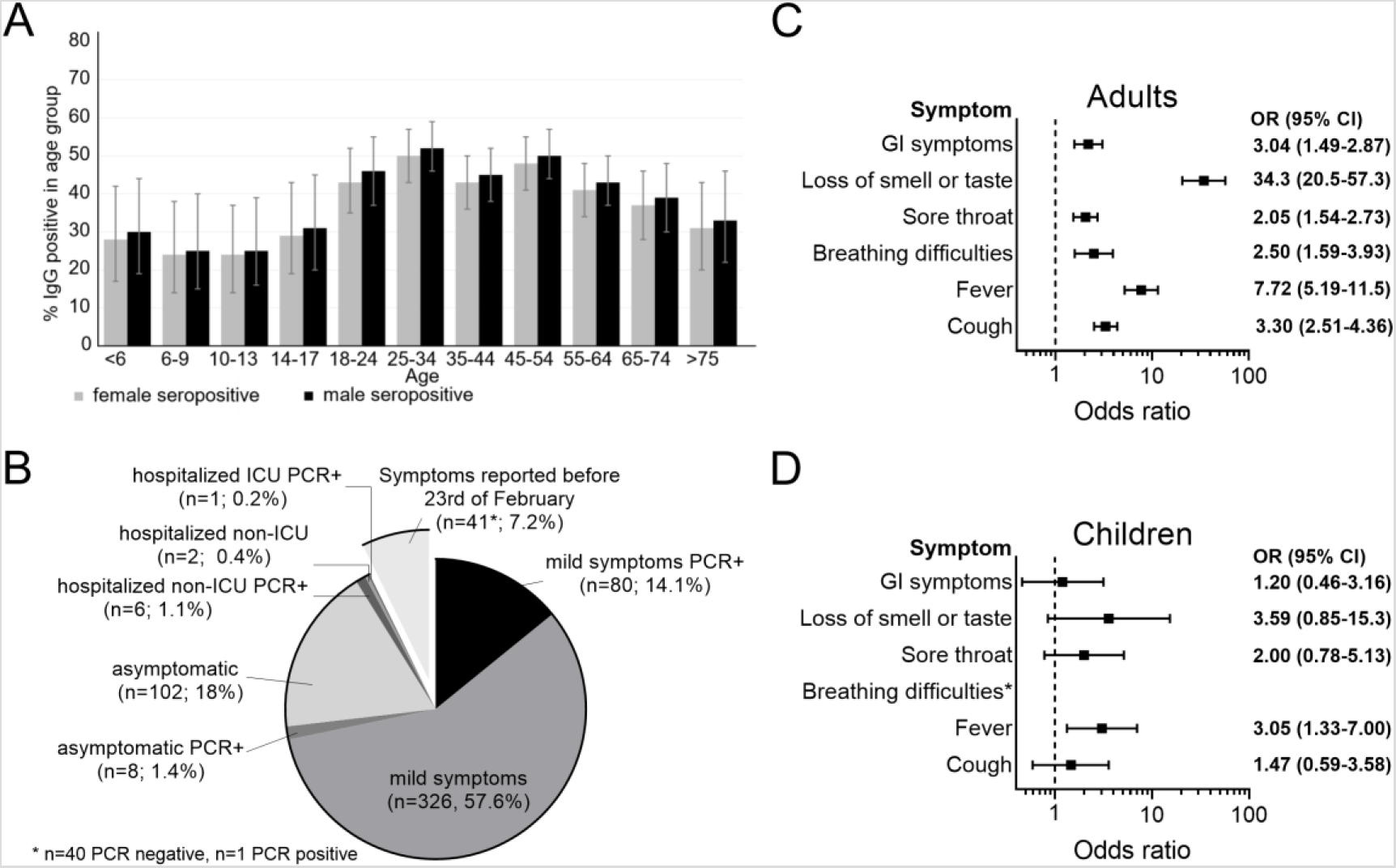
Seroprevalence and clinical course. The seroprevalence in different sex and age groups was calculated using a generalized estimating equation (GEE)-model taking household clusters into account **(A)**. Clinical courses of COVID-19 in seropositive individuals were classified based on the information provided by the study participants. Mild disease was defined as reported COVID-19-associated symptoms not requiring hospitalization. Additionally, all study participants were asked if a SARS-CoV-2 PCR was carried out and its result. Symptoms reported before February 23 were excluded as the peak of the influenza season was in the first half of February and the OR for reported symptoms and SARS-CoV-2 seropositivity were low then **(B)**. Odds ratios of self-reported symptoms regarding seropositivity in adults were calculated using generalized estimating equation (GEE)-model adjusted for age and sex, taking household clustering in account **(C)**. Odds ratios of self-reported symptoms and symptoms reported by persons of care and custody, respectively, regarding seropositivity in children were calculated using generalized estimating equation (GEE)-model adjusted for age and sex, taking household clustering in account **(D)**.

### Prevalence of virus positivity in PCR test

In 9 of the 1473 (prevalence 0.6%, 95% CI (0.3–1.2)) study participants, viral RNA was found in nasopharyngeal swabs by PCR (Table S1). In all cases, the level of viral RNA was low and all individuals presented without symptoms. Of these, 8 cases were new. Five of the 8 new cases were positive for neutralizing antibodies. Four PCR positive individuals reported to have had symptoms up to 39 days before the study of which three had the typical loss of smell and taste (Table S1). One individual had PCR confirmed COVID-19 39 days before study begin. This shows that viral RNA can be detected for longer periods even if anti-viral neutralizing antibodies are already present.

Only 105 participants reported to have been diagnosed with SARS-CoV-2 infection by PCR previously. Of these, 102 were seropositive (Table 2), showing that almost (97%) all previously PCR+ cases had seroconverted by the time the study was initiated. Thus only 102 of the 624 past SARS-CoV-2 infections diagnosed in this study by serology were previously PCR+ and reported to the authorities (16.3 %, 95% CI 13.5–19.5), corresponding to 83.7% non-diagnosed SARS-CoV-2 infections in a known hot spot considered to be the epicenter in Austria. The proportion of PCR-diagnosed and reported cases was lower in children and adolescents under the age of 18 (10.3%, 95% CI 3.9–21.2) than in adults (17.0%, 95% CI 13.9–20.3).

In Figure 2A, the results of the PCRs testing coordinated by the local authorities is given. Also, here the highest percentage of positive PCR was seen mid-March, when the highest number of participants reported to have had clinical disease.

**Figure 2.**
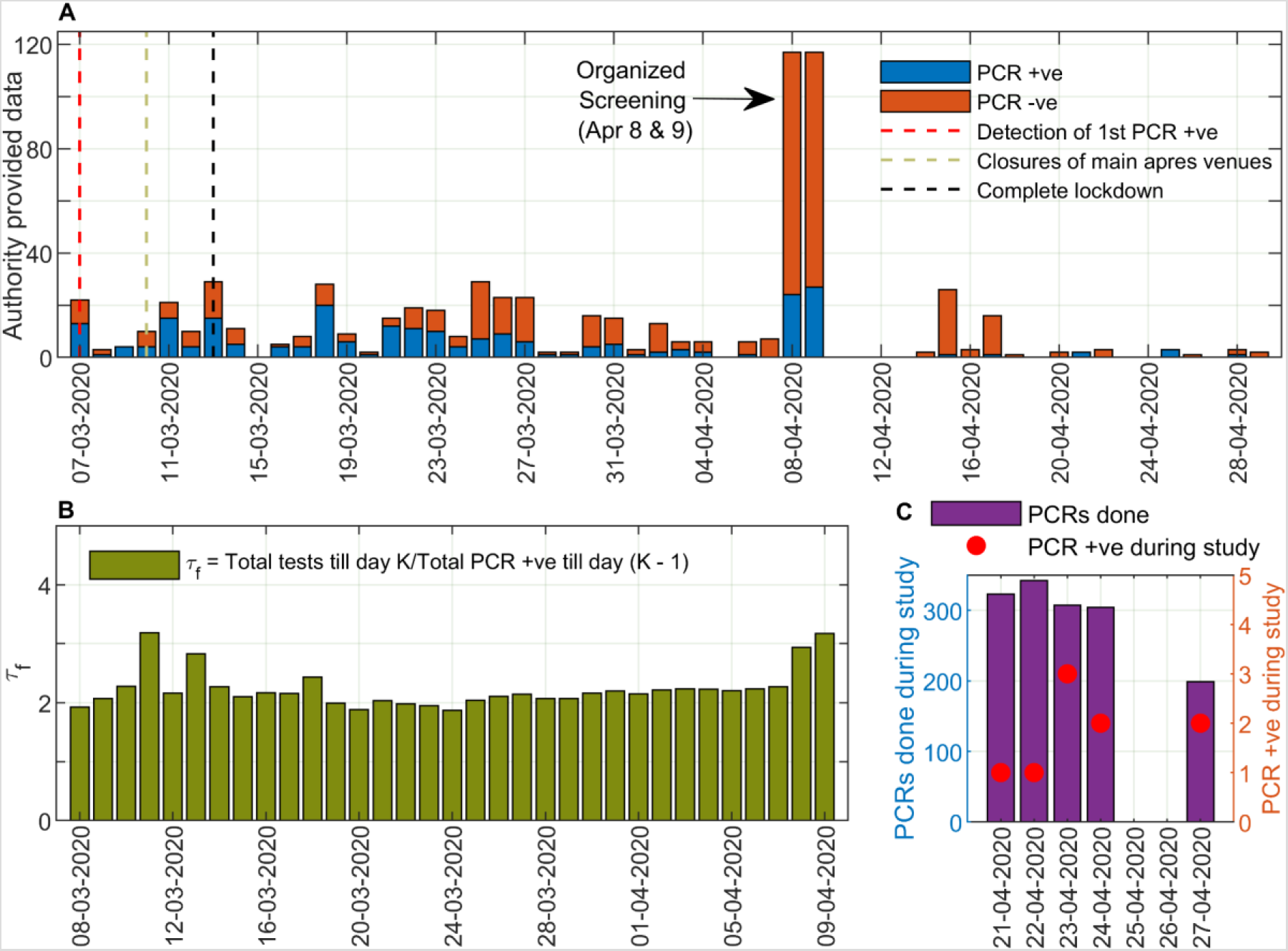
Analysis of PCR testing in Ischgl. SARS-CoV-2 PCR results from March 7 to April 29, 2020 were provided by the district administration. On March 7, 2020 the first person in Ischgl was tested positive for SARS-CoV-2. On March 10, 2020, most of the main entertainment venues in the ski resort were closed. On March 13, 2020, a complete lockdown was declared due to a rapidly increasing number of new cases and the village was quarantined. On April 8 and 9, 2020, the state health authority carried out a screening examination in which 234 people were tested for SARS-CoV-2 **(A)**. From the PCR testing data provided by the local authority, we calculated the average number of tests done per confirmed PCR result (*τ_f_*), in terms of the ratio of total PCR tests done till a day and total number of PCR positive individuals till one day prior to it **(B)**. The PCR screening during this study was performed between April 21 and April 27, 2020 in Ischgl **(C)**.

### Clinical course

For the clinical symptoms, we focused on those reported after February 23^rd^ because symptoms reported earlier were less significantly linked to SARS-CoV-2 seropositivity, most likely because the peak of the influenza season was in the first half of February and the level of SARS-CoV-2 transmission still low then. The clinical course as reported by the study participants was mild, defined as not requiring hospitalization, or even asymptomatic in most study participants, with only 9 adults (no children) reporting hospitalization (0.6%, 95% CI (0.3–1.2)) of which only one adult was in the ICU (Figure 1B). In adults, the major previous symptoms remembered by seropositive participants were typical for mild forms of COVID-19. Loss of smell and taste highly correlated with IgG positivity (OR 34.3), as did fever (OR 7.7) and cough (OR 3.3, Figure 1 C). In contrast, in the much less clinically affected children, reported previous symptoms were less significantly associated with seropositivity (Figure 1D).

Two patients in the Ischgl population died of COVID-19. The deaths both occurred after March 20. Relative to the number of seropositive individuals detected in our study, the infection fatality rate (IFR) would be 0.34%. If corrected to the number of seropositive individuals estimated to be living in Ischgl(42.4% of 1867), the IFR in Ischgl was as low as 0.25%, albeit with a broad 95% CI of 0.03–0.91. However, with only 2 deaths, this IFR in Ischgl is not statistically robust.

### Analysis of household clustering

Among 478 households, there were 184 households where all members were negative (38.5%). In 107 households, all members were positive (22.4%) and 187 households were of mixed status (39.1%). Among all 478 households, 124 households contained children. Among these households n = 84 households (67.7%) did not have any positive children, even though n = 51 of these households (60.7%) had at least one positive adult. In contrast, there was only one household among the 124 households with children where a child was positive, but no adult was positive. Mixed status households are much more common among households with children than without (Table S2). In households with children, the odds of a child of being positive were 66% lower than for adults (OR of 0.44 (95% CI 0.31–0.64).

Due to the participation rate of 79%, there may be incomplete households in this household level analysis. Thus, the estimate of the proportion of households with mixed status regarding seropositivity is conservative. In addition, there were 106 participants with unknown household information. These were excluded from the household analysis.

### Mathematical model of Ischgl outbreak

Next, we addressed with a mathematical model whether the drop in the number of new reported cases during March was solely due to the non-pharmaceutical interventions (NPIs) or whether Ischgl was on the verge of achieving herd immunity. Simulation results of the computational model (Figure MS1 - MS2) fitted well with the epidemic curve constructed with the temporal data for onset of any of the COVID-19 symptoms (Figure 3 B) and that for the onset of anosmia/dysgeusia among the seropositive individuals (Figure MS3 B) in Ischgl. As most of the residents in Ischgl were engaged in tourism related activities, one of our major assumptions was the homogeneous mixing among the residents and tourists. The basic reproduction number *R*_0_, which determines the evolution of an epidemic in a non-intervened scenario without external alterations in a local system, is meaningful at the initial phase of an outbreak before non-pharmaceutical interventions (NPIs) are introduced or behaviour of the residents has changed. Based on survey data for onset of any of the COVID-19 associated symptoms from Feb 23 2020 (Figure 3 A), we found *R*_0_ in Ischgl to be between 2.8 and 3.2, with a median value of 3.0 (Figure 3 D). This corresponds to a median value of 66.7% seroprevalence to reach herd immunity, which is consistent with herd-immunity seroprevalence calculated using a mean contact dependent daily transmission rate < *R*_1_ > for the time-windows spanning Feb 23 - Mar 16 and Feb 28 - Mar 16 (Figure 3 E).

**Figure 3.**
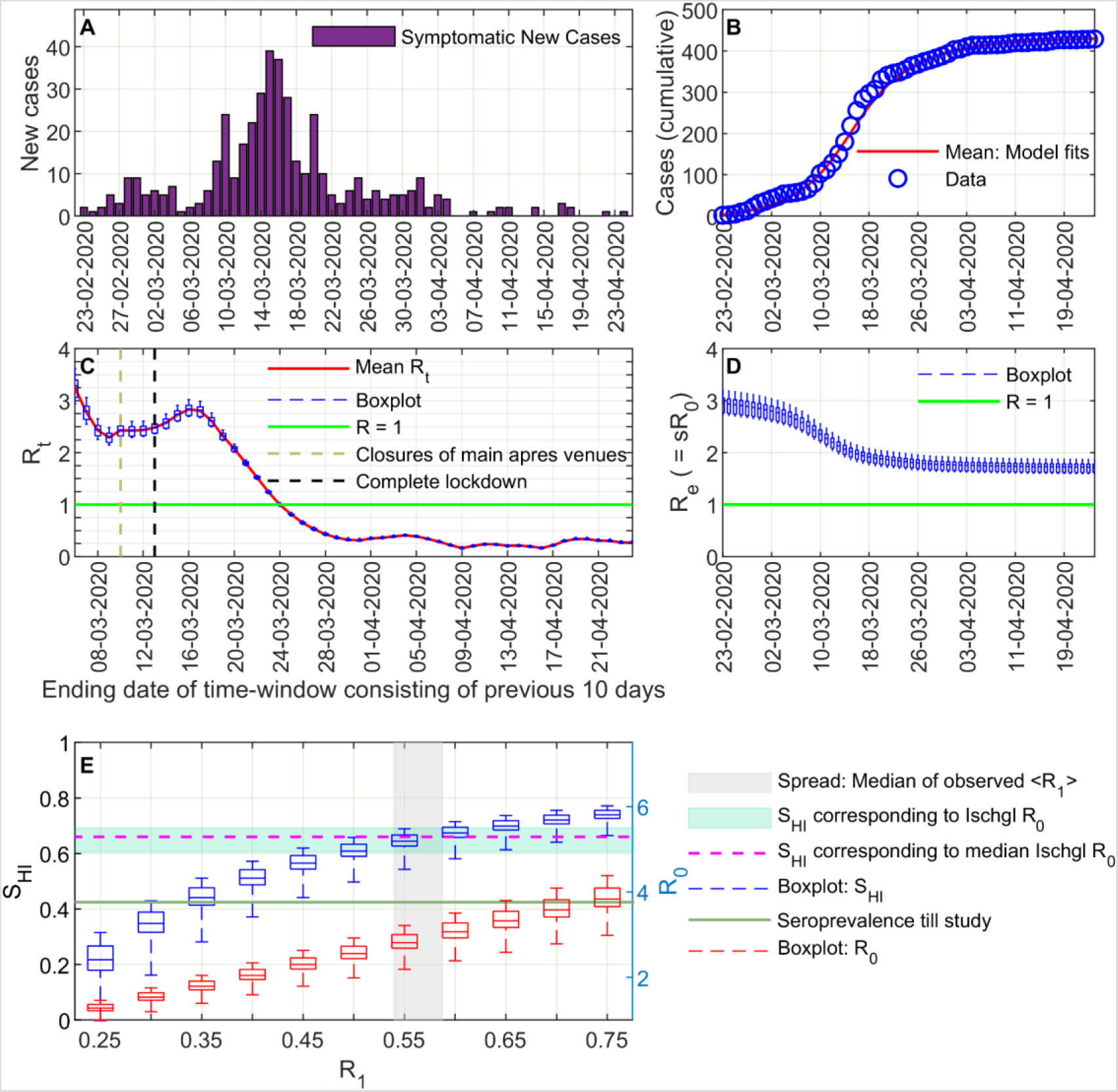
Mathematical analysis of the outbreak. The time-course of new cases in Ischgl based on any COVID-19 associated symptoms among seropositive individuals as obtained from the survey **(A)** was used to determine the model parameters (see supplementary information). The, thus, calibrated model reproduces the cumulative number of symptomatic cases **(B)** and predicts the time-dependent reproduction number *R*_t_ **(C)**. Time-dependent alterations in the effective reproduction number *R*_e_ **(D)** were calculated as *R_e_(t) = s(t) R _0_*, *s(t)* being the fraction of the susceptible population at a certain time *t*. Seroprevalence necessary for herd immunity (*S_HI_*, green shaded region) and basic reproduction number (*R*_0_, right ordinate axis, red box plots) are shown as functions of the contact dependent transmission rate (*R*_1_) **(E)**. The analysis was done with the parameter sets (except *R*_1_) best describing the case numbers until Mar 16 at a full stretch (see fitting strategy in the supplementary information). *R*_0_ in Ischgl was in between 2.8 to 3.2, with a median value of around 3.0, which corresponds to a median value of around 66.7% seroprevalence to reach herd immunity (*S_HI_* corresponding to median *R*_0_, purple line). This is consistent with herd-immunity seroprevalence (blue box plot) derived from the median contact dependent daily transmission rate < *R*_1_ > for the time-windows spanning Feb 23 - Mar 16 and Feb 28 - Mar 16 (grey patch). The actually achieved seroprevalence (green line) is substantially lower than the thresholds. Note that we considered the survey-oriented symptomatic cases before Feb 23, 2020 as asymptomatic cases of COVID-19 due to having comparably lower odds ratio (OR) than that of March 2020 and suspected overlap with the flu-season in Ischgl.

To determine the temporal variation observed during the whole course of the outbreak (Figure 3 A, 3 C), the time-dependent reproduction number was determined. We opted for a shifting time window of 10 days, in each of which *R*_t_ was determined. The next time window shifted by one day contained the history of the epidemic from the prior time window^13^ (see Supplementary Information). Ischgl faced a major outbreak till mid-March (Figure 3 C). The viral spreading infected a substantial fraction of the population between March 2 and March 18, 2020, as reflected by the drop in the effective reproduction number *R*_e_ in Figure 3-D. As *R*_e_ > 1, this implies that if all measures were lifted, a new local outbreak would be expected (Figure MS-5, MS-8). The median value of *R*_t_ was already less than 1 for the time-window ending on March 25 and afterwards (Figure 3 C). This suggests the effectiveness of the NPIs in Ischgl, which was also reflected in the projected course of the outbreak (Figure MS-6) and with different levels of reduced transmission probabilities during the period of lock-down (Figure MS-4). The same analysis based on data for anosmia/dysgeusia onset gave qualitatively similar results with a slightly higher *R*_0_ and, hence, even stronger requirements to reach herd immunity (Supplementary Figures MS3, MS7-MS9).

## Discussion

The high seroprevalence of 42.4% observed in Ischgl was based on the combination of four serological tests, two IgG immunoassays, one IgA ELISA, and a neutralizing antibody assay. The 42.4% is to our knowledge one of the highest seroprevalences reported so far.^14–18^ In New-York City a higher seroprevalence was published, but this was an analysis in patients and not the general population.^19^ In a study of the Austrian Health Ministry performed by one of Austria’s leading social research institutes (SORA) in the 27 COVID-19 hotspots in Austria outside of Tyrol, 4.6% of 269 individuals tested were seropositive.^20^ Thus, the high seroprevalence found here is in accordance with the claim that Ischgl was the epicenter of the COVID-19 pandemic in Austria and central to the spread of SARS-CoV-2 throughout Europe.

The percentage of seropositive children and adolescents under 18 (27%) was significantly lower than the percentage found in adults (45%) (OR 0.455, 95% CI 0.330–0.628, p< 0.001). This is in line with a study in Iceland^6^, Toulouse^21^, Geneva^14^ and Wuhan/Shanghai^22^, but not with other studies, where children were found to be infected as frequently as adults.^17, 23^ While the study in Wuhan/ Shanghai concluded that children are less susceptible to SARS-CoV-2 infection due to a biologic/immunological relative resistance to infection, the somewhat abrupt increase in the seroprevalence for individuals over 18 years of age found in Ischgl would argue that here the children were less exposed to the virus than the working adults. If a lower biologic susceptibility of children to SARS-CoV-2 was the only reason, the seroprevalence would be expected to already rise at younger ages and not abruptly at the age of 18, when most of the local population starts to work in local tourism. Nearly 10,000 new tourists arrived each week in addition to several 100 guests each night that come to Ischgl from other ski resorts to visit the apres-ski bars. In conclusion, the higher seroprevalence in the working population > 18 relative to children and youths is most likely at least partially explained by a higher exposure.

Of the seropositive individuals, 83.7% had not been diagnosed to have had SARS-CoV-2 infection previously. This percentage is especially high in children and asymptomatic adults and explains the less drastic difference between children and adults in our seroepidemiologic study relative to the reported PCR positive cases in Ischgl (Figure S5). The high level of unreported cases also explains the relatively low IFR of 0.25%, relative to the case fatality rate reported for Austria of 3.7%. Admittedly, the 0.25 % (0.03–0.91) IFR is not a statistically very robust number as it is based on two fatalities only. However, this low IFR is in line with a German seroepidemiologic study with an IFR of 0.37%^17^, which reported similar low numbers of fatalities, and a summary study using data from 12 seroprevalence studies (IFR 0.03–0.5%)^24^ but lower than predicted in a statistical model, 0.5 to 1.3%.^25^ This study includes all infections, and not only the reported cases. Thus, as expected, the hospitalization rate of the seropositive study participants of 1.5% was much lower than the hospitalization rates reported for Austria, which are around 15% according to official data from the Austrian registry for mandatorily reportable diseases. The low IFR and hospitalization rate could also partially by the lower seroprevalence in elderly patients. Finally, the high rate of unreported cases suggests that in future more testing especially in hotspots should be performed to allow a more efficient containment of virus spread.

The prevalence of newly PCR-positive individuals was surprisingly low and had dropped from 19% detected in a voluntary public screening of 234 individuals in Ischgl in the first week of April to 0.5% during our study 3 weeks later. This level is still higher than the 0.15% percent which dropped from a level of 0.33 % determined in the rest of Austria during the same period in a governmental surveillance study.^20,26,27^

The interesting question is now whether the achieved virus control in Ischgl is stable. Based on the basic reproduction number *R*_0_ between 2.8 and 3.2, around 66.7% seropositivity is required to stop spread of SARS-CoV-2 in Ischgl. Fitting of model parameters to the time course of case numbers in Ischgl revealed that the number of seropositive individuals in Ischgl is still substantially lower than the herd immunity threshold, thus, suggesting that the installed non-pharmaceutical measures contribute to the containment of the virus. Likely, a release of all measures leading to transmission at the same level as during the touristic season early March, would re-initiate viral spreading. However, the analysis does not allow for a precise quantitative evaluation of the herd-immunity threshold because of several limitations in the data. Subjective reporting of symptom onset dates as well as an unclear attribution of symptoms to SARS-CoV-2 infections are major sources of uncertainty. It is not known whether a fraction of the population, in particular, of the children, might combat the viral infection by innate immune responses alone without developing specific antibodies, thus, appearing seronegative in the current study. Further, the prevalence of super-spreading-events in Ischgl might impact on the mathematical analysis. Beyond our assumption of homogeneous mixing of the Ischgl residents and tourists, the precise impact of the in- and out-flow of tourists, who participated in infection chains and then left following their stay in Ischgl as well as during the exodus around March 13^th^, 2020, on the mathematical analysis cannot be assessed properly in the absence of data on the serostatus of the tourists. While the relative impact of leaving tourists and NPIs are difficult to disentangle, the drastic decline in the time-dependent reproduction number can be explained by the combination of both together with a significant immunization of the Ischgl population. The high seroprevalence alone does not explain the successful limitation of virus transmission. Population heterogeneity in terms of inherent innate and T-cell immunity, however, might further reduce the herd immunity threshold.^28,29^

One major weakness of the study is the data set generated from the structured questionnaires leading to a recall bias of PCR tests, of previous symptoms and of the associated time points. In addition, the mathematical model has several limitations that are clearly described above and in the supplement. The strengths of the study are, however, that 79% of the population in Ischgl participated including 214 children, that a very extensive antibody testing was performed that gave highly reliable results, and that one of the highest so far reported seroprevalences in a hotspot was found, which was lower in children than in adults and that was associated with a drastic decline in new infections. However, herd immunity was most likely not reached and thus the transmission rates must still be kept well under the level determined for early March 2020 to prevent virus circulation in Ischgl.

## Data Availability

Upon publication of our study, the data and codes will be available from the corresponding authors upon receipt of a suitable request.

## Acknowledgment

We thank Peter Willeit, Franz Allerberger, Bernhard Schölkopf, and Rafael Mikolajczyk for critically reading the manuscript. We thank the team of volunteers, listed in the supplement, for helping in collecting the specimen. We thank Sahamoddin Khailaie and Jesus D. B. Herrera for suggesting a technique for checking the robustness of our mathematical analysis.

## Funding

TM was supported by the Helmholtz-Association, Zukunftsthema “Immunology and Inflammation” (ZT-0027). MMH has received funding from the European Union’s Horizon 2020 research and innovation programme under grant agreement No 101003480 and the German Research Foundation (DFG) under Germany’s Excellence Strategy EXC 2155 project number 390874280. The state Tyrol funded the testing materials used in the study.

## Conflict of interest

No conflicts of interests exist. The funders had no role in the design of the study; in the collection, analyses, or interpretation of data; in the writing of the manuscript, or in the decision to publish the results.

## Notes

### Competing Interest Statement

The authors have declared no competing interest.

### Author Declarations

Ethical committee of the Medical University of Innsbruck

## References

1 Zhu, N. et al. A Novel Coronavirus from Patients with Pneumonia in China, 2019. N Engl J Med 382, 727–733, doi:10.1056/NEJMoa2001017 (2020).

2 Li, Q. et al. Early Transmission Dynamics in Wuhan, China, of Novel Coronavirus-Infected Pneumonia. N Engl J Med 382, 1199–1207, doi:10.1056/NEJMoa2001316 (2020).

3 Hoehl, S. et al. Evidence of SARS-CoV-2 Infection in Returning Travelers from Wuhan, China. N Engl J Med 382, 1278–1280, doi:10.1056/NEJMc2001899 (2020).

4 World Health Organization. (https://covid19.who.int).

5 Statistik Austria, Die Informationsmanager. (https://www.statistik.at/web_de/presse/123051.html).

6 Gudbjartsson, D. F. et al. Spread of SARS-CoV-2 in the Icelandic Population. N Engl J Med, doi:10.1056/NEJMoa2006100 (2020).

7 AGES, Österreichische Agentur für Gesundheit und Ernährungssicherheit GmbH. (https://www.ages.at/service/service-presse/pressemeldungen/ages-zur-epidemiologischen-abklaerung-des-cluster-s/), 2020).

8 Statistik Austria, Die Informationsmanager. Bevölkerung am 1.1.2019 nach Ortschaften (Gebietsstand 1.1.2019). (https://www.statistik.at/wcm/idc/idcplg?IdcService=GET_NATIVE_FILE&RevisionSelectionMethod=LatestReleased&dDocName=103419).

9 Hoffmann, M. et al. SARS-CoV-2 Cell Entry Depends on ACE2 and TMPRSS2 and Is Blocked by a Clinically Proven Protease Inhibitor. Cell 181, 271–280 e278, doi:10.1016/j.cell.2020.02.052 (2020).

10 Administration, U. S. F. D. EUA Authorized Serology Test Performance, <https://www.fda.gov/medical-devices/emergency-situations-medical-devices/eua-authorized-serology-test-performance> (

11 Clopper, C. J. & Pearson, E. S. The Use of Confidence or Fiducial Limits Illustrated in the Case of the Binomial. Biometrika Volume 26, Pages 404–413 (1934).

12 Zeger, S. L. & Liang, K. Y. Longitudinal data analysis for discrete and continuous outcomes. Biometrics 42, 121–130 (1986).

13 Khailaie, S. et al. Development of the reproduction number from coronavirus SARSCoV-2 case data in Germany and implications for political measures. PrePrint, doi:https://doi.org/10.1101/2020.04.04.20053637 (2020).

14 Stringhini, S. et al. Seroprevalence of anti-SARS-CoV-2 IgG antibodies in Geneva, Switzerland (SEROCoV-POP): a population-based study. Lancet, doi:10.1016/S0140-6736(20)31304-0 (2020).

15 Thompson C et al. Neutralising antibodies to SARS coronavirus 2 in Scottish blood donors - a pilot study of the value of serology to determine population exposure. PrePrint, doi:doi: https://doi.org/10.1101/2020.04.13.20060467 (2020).

16 Levesque J & Maybury DW. A note on COVID-19 seroprevalence studies: a meta-analysis using hierarchical modelling. PrePrint, doi:doi:10.1101/2020.05.03.20089201 (2020).

17 Streek H et al. Infection fatality rate of SARS-CoV-2 infection in a German community with a super-spreading event. PrePrint, doi:doi:10.1101/2020.05.04.20090076 (2020).

18 Sood, N. et al. Seroprevalence of SARS-CoV-2-Specific Antibodies Among Adults in Los Angeles County, California, on April 10–11, 2020 JAMA, doi:10.1001/jama.2020.8279 (2020).

19 Reifer J, Hayum N, Heszkel B, Klagsbald I & Streva VA. SARS-CoV-2 IgG Antibody Responses in New York City. PrePrint, doi:Doi: 10.1101/2020.05.23.20111427 (2020).

20 Statistik Austria, Bundesanstalt Statistik Österreich. COVID-19 Prävalenz April 2020, Ergebnisbericht. (Bundesministerium Bildung, Wissenschaft und Forschung, https://www.bmbwf.gv.at/Themen/Forschung/Aktuelles/COVID-19-Studie.html, 2020).

21 Dimeglio, C., Mansuy, J. M., Charpentier, S., Claudet, I. & Izopet, J. Children are protected against SARS-CoV-2 infection. J Clin Virol 128, 104451, doi:10.1016/j.jcv.2020.104451 (2020).

22 Zhang, J. et al. Changes in contact patterns shape the dynamics of the COVID-19 outbreak in China. Science, doi:10.1126/science.abb8001 (2020).

23 Qifang Bi, M. et al. Epidemiology and transmission of COVID-19 in 391 cases and 1286 of their close contacts in Shenzhen, China: a retrospective cohort study. The Lancet - Infectious Diseases, doi:DOI:https://doi.org/10.1016/S1473-3099(20)30287-5 (27th April, 2020).

24 Ioannidis J. The infection fatality rate of COVID-19 inferred from seroprevalence data. PrePrint, doi:doi:10.1101/2020.05.13.20101253 (2020).

25 Basu, A. Estimating the infection fatality rate among symptomatic COVID-19 cases in the United States. Health Affairs, doi:https://doi.org/10.1377/hlthaff.2020.00455 (7th of May, 2020).

26 SORA, Institute for Social Research and Consulting. Spread of SARS-CoV-2 in Austria - PCR tests in a representative sample Study report. (https://www.sora.at/fileadmin/downloads/projekte/Austria_Spread_of_SARS-CoV-2_Study_Report.pdf., 2020).

27 Statistik Austria, Die Informationsmanager. (https://www.statistik.at/web_de/presse/123051.html., 12.233–073/20).

28 Britton, T., Ball, F. & Trapman, P. A mathematical model reveals the influence of population heterogeneity on herd immunity to SARS-CoV-2. Science, doi:10.1126/science.abc6810 (2020).

29 Grifoni, A. et al. Targets of T Cell Responses to SARS-CoV-2 Coronavirus in Humans with COVID-19 Disease and Unexposed Individuals. Cell 181, 1489–1501 e1415, doi:10.1016/j.cell.2020.05.015 (2020).

